# ChatGPT for tinnitus information and support: response accuracy and retest after three months

**DOI:** 10.1101/2023.12.19.23300189

**Authors:** W. Wiktor Jedrzejczak, Piotr H. Skarzynski, Danuta Raj-Koziak, Milaine Dominici Sanfins, Stavros Hatzopoulos, Krzysztof Kochanek

## Abstract

**Background:** ChatGPT – a conversational tool based on artificial intelligence – has recently been tested on a range of topics. However most of the testing has involved broad domains of knowledge. Here we test ChatGPT’s knowledge of tinnitus, an important but specialized aspect of audiology and otolaryngology. Testing involved evaluating ChatGPT’s answers to a defined set of 10 questions on tinnitus. Furthermore, given the technology is advancing quickly, we re-evaluated the responses to the same 10 questions 3 months later.

**Material and method:** ChatGPT (free version 3.5) was asked 10 questions on tinnitus at two points of time – August 2023 and November 2023. The accuracy of the responses was rated by 6 experts using a Likert scale ranging from 1 to 5. The number of words in each response was also counted, and responses were specifically examined for whether references were provided or whether consultation with a specialist was suggested.

**Results:** Most of ChatGPT’s responses were rated as satisfactory or better. However, we did detect a few instances where the responses were not accurate and might be considered somewhat misleading. The responses from ChatGPT were quite long (averaging over 400 words) and they occasionally tended to stray off-topic. No solid references to sources of information were ever supplied, and when references were specifically asked for the sources were artificial. For most responses consultation with a specialist was suggested. It is worth noting that after 3 months the responses generally improved.

**Conclusions:** **ChatGPT pro**vided surprisingly good responses, given that the questions were quite specific. Although no potentially harmful errors were identified, some mistakes could be seen as somewhat misleading. No solid references were ever supplied. ChatGPT shows great potential if further developed by experts in specific areas, but for now it is not yet ready for serious application.

## Introduction

Chat Generative Pre-trained Transformer (ChatGPT) by OpenAI is a conversational tool based on artificial intelligence. It has recently attracted a high level of interest [1]. ChatGPT is based on large language models (LLMs) and is capable of human-like conversation. It is now being tested in various domains of knowledge including science and medicine. For example, it has been used to answer questions about national medical examinations [2,3], psychiatry [4], hemophilia [5], and colon cancer [6]. In the case of hearing, there are only a few studies that have used ChatGPT. It has been evaluated as a potential patient information source in otolaryngology [7], and has been used to assist with medical documentation in cases of Eustachian tube dysfunction [8].

In one article that has considered the future application of chatbots to hearing health care, Swanepoel and colleagues discussed its possible use by patients, clinicians, and researchers [9]. For example, in the case of patients, Swanepoel and colleagues suggest that chatbots could be used for initial screening, making recommendations for interventions, education, support, and teleaudiology. Of course, before this becomes possible, chatbots would first need to be evaluated in terms of their accuracy relative to current best knowledge.

Here we test the possible application of ChatGPT to tinnitus, an important medical and scientific topic within audiology and otolaryngology. The problem with tinnitus is that it is still not understood, especially in the case of subjective tinnitus, where the underlying pathophysiology is not known [10]. Moreover, there is no objective test [11], and there is no treatment that provides positive outcomes for the majority of sufferers [12]. At the same time, there are many sufferers: one meta-analysis reported that around 10% of the global population have chronic tinnitus [13]. Recent studies have pointed to an increase in hearing impairment as well as tinnitus, and further increases in these problems might be expected [14,15].

Kutyba and colleagues recently found that a large percentage of tinnitus sufferers actively seek out solutions for themselves [16]. We therefore assume that there might also be a considerable number of people who are inclined to turn to chatbots to learn more about their tinnitus. These could be patients, their families, students, or even physicians who want to provide the best for their patients.

The purpose of this study was to evaluate ChatGPT in terms of the accuracy of its responses to a defined set of questions about tinnitus. Furthermore, given the technology is quickly advancing, we evaluated responses to the same questions after a further 3 months.

## Material and Method

We framed 10 questions related to tinnitus, questions which in our opinion are quite common (Table 1). The questions fell into two categories. The first 5 questions were fairly basic and related to questions that might be asked by a patient. A second set of 5, more specialized, questions related to those a student of audiology or otolaryngology might ask, or someone with a deeper knowledge of tinnitus, such as a physician or researcher.

**Table 1.** Questions used to test ChatGPT.

| No | Question | Type |
| --- | --- | --- |
| 1 | I started to experience some strange sounds in the ear. What is it and what should I do? | Basic |
| 2 | How can I help myself when I suffer from tinnitus? |  |
| 3 | Should I believe in the advertisement of a medicine that treats tinnitus in one week? |  |
| 4 | How to diagnose tinnitus? |  |
| 5 | Is there a connection between hearing loss and tinnitus? |  |
| 6 | What is the difference between objective and subjective tinnitus? | Specialised |
| 7 | How the tinnitus is connected to otoacoustic emissions? |  |
| 8 | Is there link between tinnitus and psychological state of the patient? |  |
| 9 | What are the best questionnaires to evaluate tinnitus? |  |
| 10 | What is the expected result of auditory brainstem response in case of acoustic neuroma? |  |

We presented the questions to chatbot ChatGPT version 3.5 during two sessions, the first on 21.08.2023 and the second – 3 months later – on 26.11.2023. We used version 3.5 (not the paid version) since it is free and probably used by the majority of people. The responses from each session were copied to a single file (Supplementary Files 1 and 2). The set of responses was presented to 6 experts (the authors) with several years of experience in tinnitus research, documented by numerous publications. The standard of each response was rated by the experts on a 5-point Likert scale (1 = extremely unsatisfactory, 2 = unsatisfactory, 3 = neutral, 4 = satisfactory, and 5 = extremely satisfactory). Each of the experts evaluated the responses independently. Evaluations were undertaken within one week after the questions were presented to ChatGPT (at the first session, the experts were unaware of a potential follow-up session).

Additionally, the number of words in each response was counted (using Microsoft Word), whether references were provided, and whether consultation with a specialist was suggested.

Analyses were made in Matlab (version 2023b, MathWorks, Natick, MA). All datasets were tested for normality of distribution by a Lilliefors test. If the tests were passed, a t-test was then used, otherwise a nonparametric Mann–Whitney U-test. In all analyses, a 95% confidence level (p < 0.05) was taken as the criterion of significance.

## Results

Table 2 presents the average scores given to each ChatGPT response by the 6 experts for the two sessions. For 7 of the questions, the ratings of responses increased for the second session while for 3 questions they remained unchanged (one of these questions scored a maximum of 5 in both sessions). However all changes were not statistically significant.

**Table 2.**
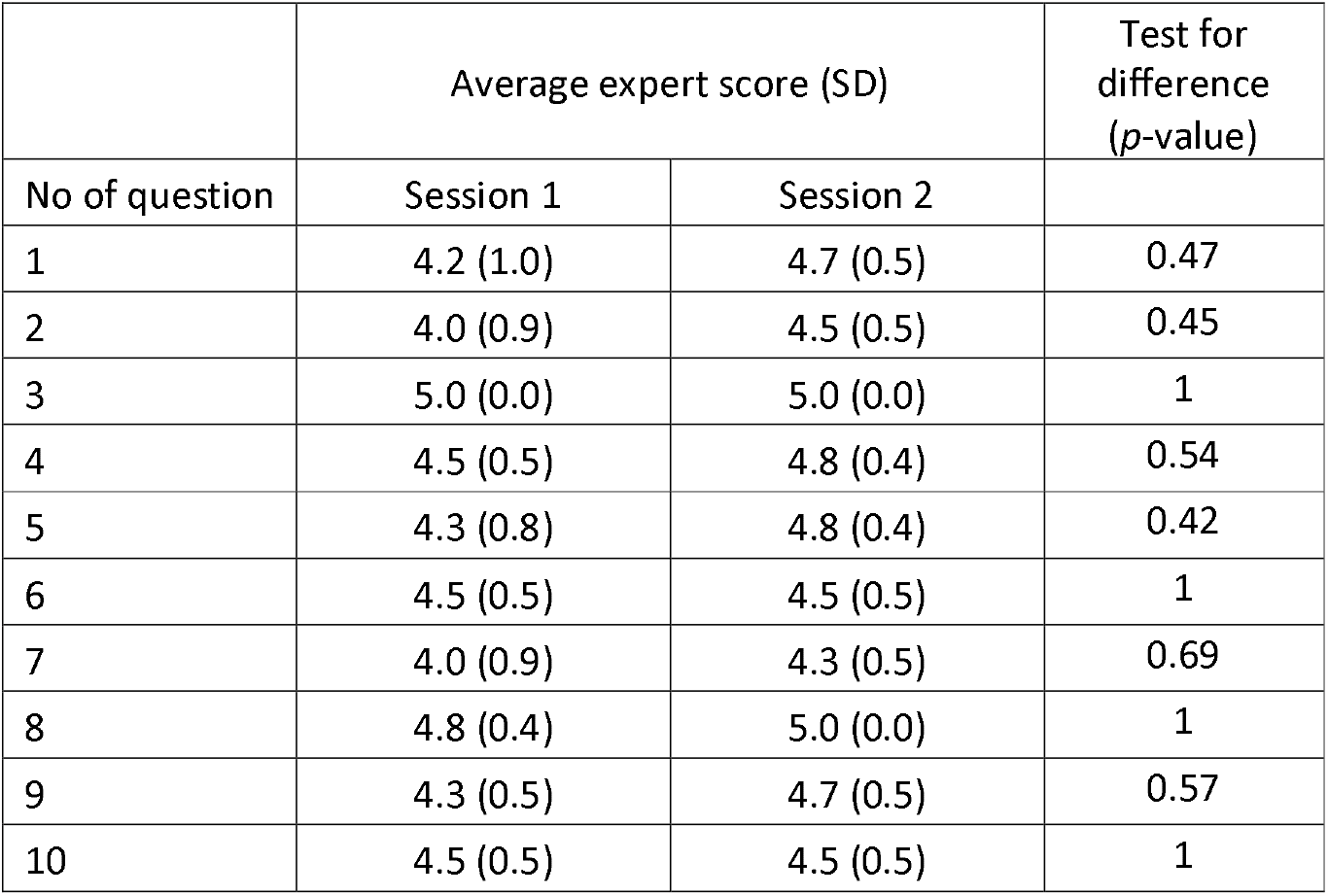
Average expert scores for each response provided by ChatGPT at the two sessions (session 1 at August 2023 and session 2 at November 2023). Results of statistical comparison between sessions are provided in the last column. (SD – standard deviation).

The difficulty of the questions did not change the average rating. There were no statistically significant differences between the scores for basic and specialized responses.

Table 3 shows the overall average for all questions, and the number of responses that were either satisfactory or extremely satisfactory. The average expert score (average of ratings for responses to questions 1–10) increased significantly for the second session. Similarly, there was a significant increase between the sessions irrespective of whether the questions were basic or specialized.

**Table 3.** Total average score given by the experts and number of responses rated as 5 (extremely satisfactory) and 4 and 5 (satisfactory and extremely satisfactory together). Results of statistical comparison between sessions are provided in the last column. Significant differences marked by asterisks. SD – standard deviation.

|  | Mean (SD) |  | Test for difference<br>(*p*-value) |
| --- | --- | --- | --- |
|  | Session 1 | Session 2 |  |
| Total average expert score (max = 5) | 4.4 (0.3) | 4.7 (0.2) | 0.0031* |
| Average expert score – basic (max = 5) | 4.4 (0.8) | 4.8 (0.4) | 0.0028* |
| Average expert score – specialized (max = 5) | 4.4 (0.6) | 4.6 (0.5) | 0.023* |
| Average number of responses rated as 5 (max = 10) | 5.3 (2.3) | 6.8 (2.4) | 0.017* |
| Average number of responses rated as 4 or better (max = 10) | 8.8 (1.5) | 10 (0.0) | 0.109 |

It can be seen that most of the responses given by ChatGPT were rated as satisfactory or better in the first session and all of them in the second (Table 3). For the second session there was a significant increase in the number of responses rated as 5. Nevertheless, there were a few instances where the responses were not perfectly true and might be considered slightly misleading (especially for the first session). This aspect is further explored in the Discussion.

The scores assigned by each expert to ChatGPT’s responses were compared to a mid-rating of 3 (i.e. a total score of 30 for 10 responses). Our expectation was that a competent set of answers should achieve a better rating than that. Despite some differences between the experts, all scores were significantly better than the mid rating for the first session as well as for the second (*p* < 0.0001).

Responses given by ChatGPT were quite long, on average 431 words for the first session and 411 for the second (Table 4). All responses lacked references to sources of information, and when specifically asked for references the tool generated artificial ones. This behavior is described in more detail in the discussion. For most responses there was a suggestion to consult a specialist for the first session while in the second session all the responses contained such a suggestion (Table 4).

**Table 4.** Number of ChatGPT responses that provided sources of information, number that suggested the help of a specialist, and the average number of words in a response at the two sessions. Results of statistical comparison between sessions are provided in the last column. SD – standard deviation.

|  | Session 1 | Session 2 | Test for difference (p-value) |
| --- | --- | --- | --- |
| Number of responses in which sources were provided (0/1, max = 10) | 0 | 0 | – |
| Number of responses in which the help of a specialist was suggested (0/1, max = 10) | 8 | 10 | 0.13 |
| Average number of words (SD) | 431 (46) | 411 (40) | 0.065 |

## Discussion

The majority of ChatGPT responses were rated as satisfactory or better. There were only a few scores which fell below this level. This was the case only in the first session, and it seems that the quality of responses improved significantly over the intervening 3 months. We were somewhat surprised at the level of competence, as tinnitus is a complex topic and many aspects cannot be clearly explained. In our opinion, ChatGPT provides perfectly reasonable responses, especially to basic questions. The answers were in general easy to read and understand, and were quite comprehensive. They were usually clearer than the responses we typically get from most of our students. One important aspect, and one we regard positively, is that ChatGPT conveys the importance of professional assessment and care in cases of tinnitus. Furthermore, it reinforces that treatment depends on the needs of each patient and, to be highly effective, often needs to encompass multiple approaches.

At the same time, ChatGPT’s responses sometimes lacked a degree of focus. They often consisted of a list of topics related to the question but without any detailed analysis, clear connections, or deeper aspects of a problem. This seemed to reflect the fact that most of the responses were too long and contained extraneous information. Sometimes ChatGPT appeared to add extra information just to get to a predetermined number of words.

When we looked at actual responses, certain patterns of successes and failures began to emerge. This was especially the case in the first session (the reader can explore the responses themselves in supplementary files 1 and 2). Starting with the positives, we draw attention to question 3, which was constructed specifically to address the problem of advertisements which promise quick remedies for tinnitus. These advertisements are commonly encountered on web pages, news portals, or are sent by email. They are potentially harmful in that, as well as losing money, patients may put off contacting a specialist and delay proper treatment. The ChatGPT response in this case is very balanced. It starts with saying “It’s important to approach advertisements for medications that claim to treat tinnitus in a very short period of time with caution and skepticism.” It continues by giving more detail about the complexity of tinnitus, the need for scientific support, and placebo effects (see Supplementary file 1), and finishes with a recommendation about seeking personalised advice from a healthcare professional. In our opinion, this response shows the great potential of chatbots to provide reasonable answers and recommendations about difficult and controversial issues.

We now move on to examples which show the sort of mistakes ChatGPT can make. In response to questions 1 and 2, ChatGPT said that reducing caffeine and salt intake may reduce tinnitus symptoms. In fact, although there are some studies pointing in this direction, the subject is controversial and there is certainly no definite proof [17].

Question 7 asked about the relation of tinnitus to otoacoustic emissions (OAEs). It replied only about evoked OAEs, even though OAEs can also be spontaneous [18] and these can be said to most closely resemble tinnitus. However, ChatGPT did not state that fact, mentioning only evoked OAEs and not giving any indication about different types of OAEs. There are some reports of spontaneous OAEs that have the same frequency as the subject’s tinnitus [19], and this is highly relevant. However during session 1, the response only said, “For instance, individuals with tinnitus that is predominantly tonal might exhibit different OAE patterns compared to those with non-tonal tinnitus.” This is too general and needs further detail. Without additional information about the type of OAE (evoked or spontaneous), the measurement paradigm, and the OAE parameters that are relevant, this sentence is essentially useless. Moreover, the whole answer was quite long and convoluted, compounded by a lack of references to specific information. In session 2 there seemed to be only a small improvement in this response.

Question 8 related to a link between tinnitus and the psychological state of the patient. Although the response was generally correct, it does not mention that such a link is usually present only for cases of severe chronic tinnitus, not for all instances of tinnitus [20].

Question 9 asked about questionnaires used to study tinnitus. The response for session 1 mentioned a few popular questionnaires like the Tinnitus Handicap Inventory (THI) [21], the Tinnitus Functional Index (TFI) [22], and also, remarkably, a “Goebel and Hiller Questionnaire”. But we were unable to find such a thing in the literature. Goebel and Hiller did contribute to studies based on some questionnaires, but there seems to be no actual questionnaire named after them [23,24]. This seems to be a creation of ChatGPT, creations that are sometimes called ‘chatbot hallucinations’ [25]. Fortunately, this problem was not present in the response during the second session, which also listed a smaller number of questionnaires. On the other hand, there was no mention of some newer questionnaires that are gaining attention (e.g. [26]). Also ChatGPT said nothing about the need to have questionnaires adapted to the patient’s native language.

The major drawback of ChatGPT is that, when not directly asked, it does not provide references. Such a lack may not be needed in casual or fun conversation, but it is essential when discussing scientific or medical knowledge. When the phrase “with references” was added to each question, ChatGPT provided them during the first session, but they were totally artificial. False citations were created, with some plausible author names, some plausible title, and some journal in which papers on the topic are published. This is similar to the creations or hallucinations mentioned in connection with the “Goebel and Hiller Questionnaire”. During the second session we observed a change in that direct links to references were provided together with a title (see Fig. 1). Unfortunately, these too were fake. A screenshot of a portion of the response to question 8 shows four references. The first directs to a page which says: “Sorry! That page doesn’t seem to exist”. The second and fourth link to scientific papers on topics not related to tinnitus and with titles different to those ChatGPT gives. The third links to a paper related to tinnitus but with a title once again different to that in ChatGPT’s response.

**Figure 1.**
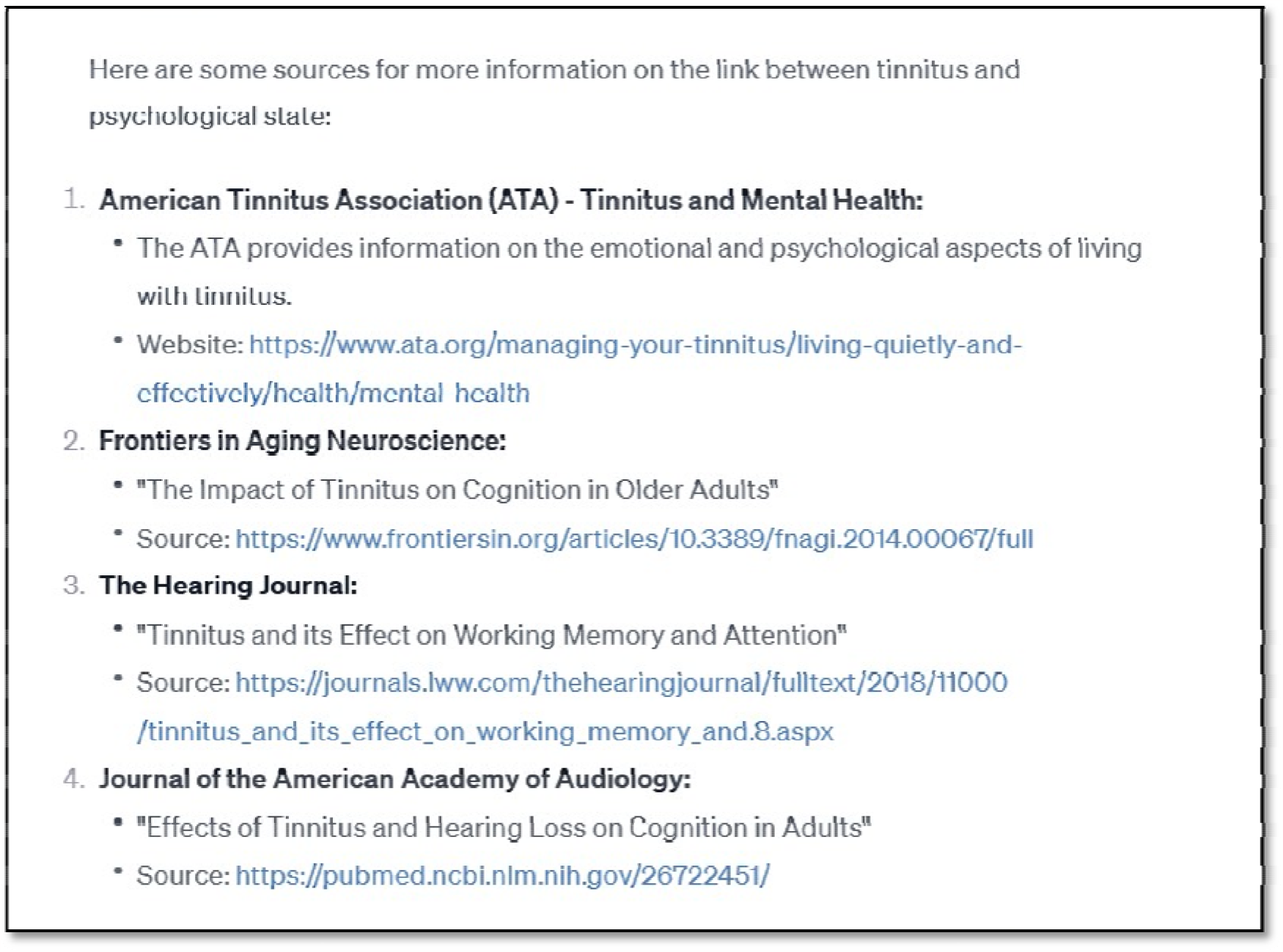
A screenshot of a portion of a ChatGPT response to question number 8 in which references were asked for. The prompt used was: “Is there link between tinnitus and psychological state of the patient? Provide sources of information.” Only the part of the response with references is shown.

Often, in response to a specialised question, ChatGPT will provide a reasonable disclaimer, suggesting that a specialist should be consulted, or that scientific knowledge is progressing and information might be out of date. A few times it said that it was powered by knowledge up to September 2021. We view this aspect as a positive, since not many sources will suggest you should consult a specialist or that knowledge is limited (even if it is suggested, it is usually only as part of a reference or as a disclaimer in small font). With ChatGPT, it was included as part of the whole response.

Finally, we must remember that ChatGPT was created to provide intelligent content to open questions, not to be a perfect scientific or educational tool. Taking this into consideration, we are generally impressed with its capabilities.

## Conclusions

On our set of 10 questions, ChatGPT provided surprisingly good responses, especially given the fact that the questions were on a very specific topic. Although no potentially harmful errors were identified, there were some omissions that could be considered somewhat misleading. Scientifically, the big drawback is that no references were ever provided, and when specifically asked to provide references they were artificial. ChatGPT shows great potential if developed further by specialists in a particular area, but for now it is not yet ready to be used for serious applications. On the positive side, it seems that over the 3 months of our study the response quality improved significantly.

## Data Availability

All data produced in the present work are contained in the manuscript and supplementary files.

