## Supplementary File 1 for "ChatGPT for tinnitus information and support: response accuracy and retest after three months"

Responses by ChatGPT – 21.08.2023

**1.**

I started to experience some strange sounds in the ear? What is it and what should I do?

ChatGPT

Experiencing strange sounds in the ear, such as ringing, buzzing, hissing, or other noises, is a common symptom of a condition known as tinnitus. Tinnitus is the perception of sound in the absence of any external sound source. It can affect one or both ears and may vary in intensity and nature. Tinnitus can be temporary or chronic and is often associated with hearing loss, exposure to loud noise, ear infections, or other underlying health issues.

If you're experiencing tinnitus, here are some steps you can take:

1. **Consult a Healthcare Professional:** It's important to consult with a healthcare professional, such as an audiologist or an ear, nose, and throat (ENT) specialist. They can perform a thorough evaluation to determine the underlying cause of your tinnitus and recommend appropriate treatment options.
2. **Medical History and Examination:** The healthcare provider will likely ask about your medical history, any recent exposure to loud noises, medications you're taking, and any other relevant information. They will also conduct a physical examination of your ears.
3. **Audiological Evaluation:** An audiologist may perform hearing tests to assess your hearing and determine if there's any hearing loss associated with your tinnitus.
4. **Identify Underlying Causes:** Tinnitus can be a symptom of various underlying conditions, including hearing loss, ear infections, earwax buildup, temporomandibular joint (TMJ) disorders, and more. Identifying and addressing the underlying cause can help manage the tinnitus.
5. **Treatment Options:** The treatment for tinnitus depends on its underlying cause. Some potential treatment options include:
   - **Hearing Aids:** If hearing loss is contributing to your tinnitus, hearing aids can help by improving your overall hearing and reducing the perception of tinnitus.
   - **Sound Therapy:** This involves using external sounds, such as white noise, music, or nature sounds, to distract from and reduce the perception of tinnitus.
   - **Counseling and Cognitive Behavioral Therapy (CBT):** These approaches can help you cope with the emotional distress and anxiety often associated with tinnitus.
   - **Tinnitus Retraining Therapy (TRT):** TRT combines sound therapy and counseling to help individuals habituate to the perception of tinnitus.
   - **Medications:** In some cases, medications such as antidepressants or antianxiety drugs may be prescribed to help manage the emotional aspects of tinnitus.
   - **Lifestyle Changes:** Avoiding loud noises, managing stress, maintaining good ear hygiene, and reducing caffeine and salt intake can also contribute to tinnitus management.
6. **Follow Recommendations:** If a healthcare professional provides recommendations for managing your tinnitus, make sure to follow them closely. Tinnitus management may require time and patience, and a combination of approaches might be necessary for effective relief.

Remember, it's crucial to seek professional advice if you're experiencing tinnitus, as the underlying cause could vary widely. Avoid self-diagnosis and instead consult with a qualified healthcare provider to receive accurate assessment and guidance tailored to your situation.

**2.**

How can I help myself when I suffer from tinnitus?

ChatGPT

If you're experiencing tinnitus, there are several strategies you can try to help manage the condition and alleviate its impact on your daily life. Keep in mind that the effectiveness of these strategies can vary from person to person, so it's a good idea to experiment and see what works best for you. Here are some self-help tips for managing tinnitus:

1. **Relaxation Techniques:**
   - Practicing relaxation techniques such as deep breathing, meditation, yoga, or progressive muscle relaxation can help reduce stress and anxiety, which can exacerbate tinnitus.
2. **Sound Therapy:**
   - Background noise or sound therapy can help mask the perception of tinnitus. You can use devices that generate white noise, pink noise, or nature sounds. Experiment with different types of sounds to find what works for you.
3. **Avoid Loud Noises:**
   - Protect your ears from loud noises, as exposure to loud sounds can worsen tinnitus. Use earplugs or earmuffs in noisy environments and turn down the volume on electronic devices.
4. **Healthy Lifestyle:**
   - Maintain a healthy lifestyle by eating a balanced diet, staying hydrated, and getting regular exercise. These factors can contribute to overall well-being, which may indirectly help with tinnitus management.
5. **Reduce Stress and Anxiety:**
   - Stress and anxiety can exacerbate tinnitus. Engage in activities you enjoy, practice relaxation techniques, and consider counseling or therapy to manage emotional distress.
6. **Limit Caffeine and Salt:**
   - Some people find that reducing their caffeine and salt intake can help improve their tinnitus symptoms. Experiment with your diet to see if there's a positive effect.
7. **Avoid Silence:**
   - Complete silence can make tinnitus more noticeable. Having some background noise, such as a fan or soft music, can help divert your attention away from the tinnitus sounds.
8. **Avoid Alcohol and Smoking:**
   - Alcohol and smoking can affect blood flow and worsen tinnitus in some cases. Reducing or eliminating these substances may help alleviate symptoms.
9. **Get Adequate Sleep:**
   - Ensure you're getting enough restful sleep. Establish a regular sleep routine, create a comfortable sleep environment, and consider using white noise or relaxation techniques to help you sleep better.
10. **Support Groups and Counseling:**
    - Joining a tinnitus support group or seeking counseling can provide you with a space to share your experiences, learn from others, and receive emotional support.
11. **Stay Informed:**
    - Educate yourself about tinnitus. Understanding the condition and its possible causes can help you better manage your symptoms and make informed decisions about treatment options.
12. **Consult a Healthcare Professional:**
    - If your tinnitus is causing significant distress or interfering with your daily life, it's important to consult a healthcare professional, such as an audiologist or ENT specialist. They can provide personalized guidance and recommend appropriate treatments based on your individual situation.

Remember that tinnitus management can be a gradual process, and what works for one person might not work for another. Be patient with yourself and continue to explore different strategies until you find a combination that helps improve your quality of life.

**3.**

Should I believe in the advertisement of a medicine that treats tinnitus in one week?

ChatGPT

It's important to approach advertisements for medications that claim to treat tinnitus in a very short period of time with caution and skepticism. While there have been advancements in tinnitus research and treatment, it's unlikely that a legitimate and effective treatment for tinnitus would work for everyone within just one week. Here are a few reasons to be cautious:

1. **Complexity of Tinnitus:** Tinnitus is a complex condition with various underlying causes and contributing factors. What works for one person might not work for another due to the diversity of causes and individual responses.
2. **Lack of Scientific Support:** Many claims made in advertisements for quick-fix tinnitus treatments may not be supported by rigorous scientific research or clinical trials. Validating the safety and efficacy of a treatment typically requires extensive testing and peer-reviewed studies.
3. **Personalized Treatment:** Effective tinnitus management often involves a personalized approach based on an individual's specific symptoms, causes, and medical history. There is no one-size-fits-all solution.
4. **Placebo Effect:** Some treatments that promise rapid relief may work due to the placebo effect, where a person's belief in the treatment influences their perception of improvement. This effect can be temporary and might not address the root cause of tinnitus.
5. **Risk of Scams:** Unfortunately, the field of tinnitus treatment is not immune to scams or fraudulent products that prey on people's desperation for relief. Be cautious of products with limited or no credible information backing their claims.
6. **Consultation with Healthcare Professionals:** It's always advisable to consult with a qualified healthcare professional, such as an audiologist or ENT specialist, before trying any new treatment. They can provide evidence-based recommendations tailored to your specific situation.

When considering a new treatment, especially one that promises quick results, here are some steps you can take:

- Research the treatment and its manufacturer to see if there's credible scientific evidence or reputable testimonials supporting its effectiveness.
- Consult with a healthcare professional to discuss the treatment's claims and evaluate its suitability for your condition.
- Be wary of treatments that require significant upfront payments or subscription fees without clear evidence of effectiveness.
- Remember that the best approach to managing tinnitus involves evidence-based treatments, lifestyle adjustments, and guidance from qualified medical professionals.

In summary, while advancements are being made in tinnitus research and treatment, it's important to be cautious when encountering advertisements that promise rapid relief. Always prioritize treatments with a solid foundation of scientific research and consult with healthcare professionals for personalized advice.

**4.**

How to diagnose tinnitus?

ChatGPT

Diagnosing tinnitus involves a comprehensive assessment that includes a medical history interview, physical examination, and often specialized audiological evaluations. The goal is to determine the underlying cause of the tinnitus and its impact on the individual. Here's an overview of the diagnostic process for tinnitus:

1. **Medical History:** The healthcare provider will begin by asking you questions about your medical history, including:
   - When you first noticed the tinnitus.
   - The nature of the sounds you're hearing (e.g., ringing, buzzing, hissing).
   - Whether the tinnitus is constant or intermittent.
   - Any associated symptoms, such as hearing loss, dizziness, or ear pain.
   - Any recent exposure to loud noise or traumatic events.
2. **Physical Examination:** A physical examination of your ears, head, neck, and sometimes the jaw (temporomandibular joint) may be performed to check for any visible signs of earwax buildup, infection, inflammation, or other potential causes of tinnitus.
3. **Audiological Evaluation:** An audiologist will conduct a series of tests to assess your hearing and gather information about your tinnitus. These tests may include:
   - **Pure-tone audiometry:** This measures your hearing thresholds at different frequencies and can determine if there's any hearing loss present.
   - **Speech audiometry:** This evaluates your ability to understand speech at different volume levels.
   - **Tympanometry:** This assesses the function of the middle ear and the mobility of the eardrum.
   - **Otoacoustic emissions (OAEs):** This test checks the function of the outer hair cells in the inner ear.
4. **Tinnitus Assessment:** During the audiological evaluation, the audiologist will also assess the characteristics of your tinnitus, such as:
   - Loudness and pitch of the tinnitus sounds.
   - Whether the tinnitus is localized to one ear or appears to be centralized in the head.
   - The impact of tinnitus on your daily life and emotional well-being.
5. **Imaging and Additional Tests:** Depending on the findings and suspected causes, additional tests such as magnetic resonance imaging (MRI), computed tomography (CT) scans, or blood tests may be recommended to rule out more serious underlying conditions.
6. **Consultation with Specialists:** If necessary, your healthcare provider may refer you to an ear, nose, and throat (ENT) specialist or other medical specialists to further investigate the cause of your tinnitus.
7. **Diagnosis and Treatment Plan:** Once the evaluation is complete, your healthcare provider will discuss the findings with you and provide a diagnosis. The diagnosis will guide the development of a personalized treatment plan based on the underlying cause of your tinnitus.

It's important to note that tinnitus can have various underlying causes, ranging from exposure to loud noise to certain medical conditions. Because of this complexity, accurate diagnosis requires a thorough evaluation by qualified healthcare professionals. If you're experiencing tinnitus, it's advisable to seek medical attention and follow the guidance of your healthcare provider.

**5.**

Is there a connection between hearing loss and tinnitus?

ChatGPT

Yes, there is a strong connection between hearing loss and tinnitus. Many individuals with tinnitus also have some degree of hearing loss, and the two conditions often coexist. Here's how hearing loss and tinnitus are related:

1. **Shared Causes:** Both hearing loss and tinnitus can be caused by similar factors. Exposure to loud noises, aging, certain medical conditions (such as Meniere's disease), ear infections, and ototoxic medications are examples of factors that can contribute to both hearing loss and tinnitus.
2. **Cochlear Damage:** The inner ear (cochlea) plays a crucial role in both hearing and the perception of tinnitus. Damage to the hair cells within the cochlea, often due to noise exposure or aging, can lead to both reduced hearing sensitivity and the perception of tinnitus.
3. **Compensatory Mechanisms:** When the hair cells in the inner ear are damaged, the brain might attempt to compensate for the lack of incoming sound by increasing its neural activity. This increased activity can contribute to the perception of tinnitus.
4. **Brain Plasticity:** The brain's response to reduced auditory input due to hearing loss can involve changes in neural pathways and increased sensitivity to neural activity. This neuroplasticity can contribute to the perception of tinnitus sounds even when there's no external auditory stimulation.
5. **Tinnitus as a Symptom:** In some cases, tinnitus can be a symptom of underlying hearing loss. As the brain tries to compensate for reduced auditory input, it might generate phantom sounds, leading to the perception of tinnitus.
6. **Treatment Interactions:** Treating hearing loss can sometimes also lead to improvements in tinnitus. For example, using hearing aids to amplify external sounds can help mask tinnitus and provide the brain with more auditory input, which can reduce the perception of tinnitus.

It's important to note that while hearing loss and tinnitus often coexist, not everyone with hearing loss will experience tinnitus, and vice versa. Additionally, the severity of both conditions can vary widely from person to person.

If you're experiencing either hearing loss or tinnitus, it's advisable to consult with a healthcare professional, such as an audiologist or ENT specialist. They can evaluate your condition, recommend appropriate tests, and provide personalized guidance on managing both hearing loss and tinnitus. Addressing hearing loss through appropriate interventions, such as hearing aids or other assistive devices, can often lead to improved overall auditory perception and quality of life.

**6.**

What is the difference between objective and subjective tinnitus?

ChatGPT

Objective tinnitus and subjective tinnitus are two different classifications of tinnitus based on how the perception of the sound is experienced and detected. Here's the difference between the two:

1. **Objective Tinnitus:** Objective tinnitus refers to a type of tinnitus where the perceived sound can be heard not only by the person experiencing it but also by a healthcare professional during a physical examination. This type of tinnitus is relatively rare and is often associated with a physical source or a vascular problem. Some common causes of objective tinnitus include:
   - Muscle spasms: Abnormal muscle contractions in the ear or surrounding areas can produce sounds that others can hear.
   - Vascular abnormalities: Blood flow turbulence or changes in blood vessels near the ear can create audible sounds.
   - Patulous Eustachian tube: A dysfunction in the Eustachian tube can lead to abnormal sounds related to breathing or swallowing.

Objective tinnitus can sometimes be identified and localized through careful examination by a healthcare professional.

2. **Subjective Tinnitus:** Subjective tinnitus is the more common type of tinnitus and refers to the perception of sound in the absence of any external sound source. This perception is solely experienced by the person with tinnitus and cannot be heard by others. The sounds associated with subjective tinnitus can vary widely and may include ringing, buzzing, hissing, whistling, clicking, or other types of noises.

Subjective tinnitus is often linked to factors like hearing loss, noise exposure, ear infections, or various underlying health conditions. It can be challenging to pinpoint the exact cause of subjective tinnitus, and its perception can be influenced by factors such as stress, anxiety, and attention.

In summary, the key difference between objective and subjective tinnitus lies in whether the perceived sound is audible to others. Objective tinnitus is a rare form in which the sound can be heard by both the affected individual and a healthcare professional, often indicating a specific physical cause. Subjective tinnitus, on the other hand, is the more common type where only the person experiencing it can hear the sounds, and it is often associated with various factors related to hearing and the auditory system.

**7.**

How the tinnitus is connected to otoacoustic emissions?

ChatGPT

Otoacoustic emissions (OAEs) are sounds that are generated by the inner ear (specifically, the cochlea) in response to external auditory stimuli. These emissions can be measured using sensitive microphones placed in the ear canal. OAEs are commonly used to assess the health and function of the outer hair cells in the cochlea, which play a crucial role in the process of hearing.

The connection between tinnitus and otoacoustic emissions lies in their association with cochlear health and function. Here's how tinnitus and OAEs are connected:

1. **Cochlear Health:** OAEs are a valuable tool for evaluating the integrity and health of the cochlea, particularly the outer hair cells. Outer hair cells are responsible for amplifying and fine-tuning auditory signals, which is essential for normal hearing sensitivity. Damage to these cells, often caused by factors like noise exposure or aging, can lead to hearing loss and potentially contribute to the perception of tinnitus.
2. **Tinnitus and Cochlear Damage:** Many cases of tinnitus are associated with cochlear damage, including damage to the outer hair cells. As outer hair cells play a role in amplifying sound signals, their dysfunction can lead to reduced auditory input to the brain. This reduction in normal auditory input is believed to contribute to the brain's generation of phantom sounds, resulting in the perception of tinnitus.
3. **OAEs and Tinnitus Studies:** Researchers have conducted studies to explore the relationship between OAEs and tinnitus. Some studies have found that individuals with tinnitus may exhibit changes in OAE responses compared to those without tinnitus. These changes can include alterations in OAE amplitudes, latencies, or other characteristics.
4. **Tinnitus Subtypes and OAEs:** Research has also suggested that different subtypes of tinnitus might be associated with specific patterns of OAE changes. For instance, individuals with tinnitus that is predominantly tonal might exhibit different OAE patterns compared to those with non-tonal tinnitus.
5. **Clinical Application:** In clinical practice, OAEs can be used as part of the assessment process for individuals with tinnitus. Changes in OAE responses might indicate cochlear dysfunction that could be contributing to the tinnitus perception. OAE measurements can provide valuable information about the status of the cochlea and the outer hair cells, which can guide treatment decisions.

While the connection between tinnitus and OAEs is an active area of research, it's important to note that not all cases of tinnitus will have associated changes in OAEs, and the relationship can vary among individuals. If you're experiencing tinnitus, consulting with a healthcare professional, such as an audiologist, can help determine whether OAE testing is appropriate as part of your comprehensive evaluation.

**8.**

Is there link between tinnitus and psychological state of the patient?

ChatGPT

Yes, there is a strong link between tinnitus and the psychological state of the patient. Tinnitus can have a significant impact on a person's emotional well-being and overall psychological health. The experience of tinnitus can lead to various psychological reactions, and conversely, a person's psychological state can influence how they perceive and cope with tinnitus. Here are some ways in which tinnitus and psychological well-being are connected:

1. **Emotional Distress:** Tinnitus can lead to a range of negative emotions, including anxiety, depression, frustration, irritability, and even feelings of hopelessness. The persistent presence of an internal sound that cannot be escaped can be distressing for many individuals.
2. **Anxiety and Stress:** Tinnitus can contribute to heightened levels of anxiety and stress. Individuals may become preoccupied with the sound, fearing that it will worsen or that it signifies a more serious underlying condition. This preoccupation can lead to chronic stress and anxiety.
3. **Depression:** The emotional burden of living with tinnitus can contribute to the development or exacerbation of depression. Sleep disturbances often associated with tinnitus can further contribute to depressive symptoms.
4. **Attention and Focus:** Tinnitus can capture a person's attention, making it difficult to concentrate on tasks or enjoy activities. This shift in focus towards the tinnitus sound can lead to reduced engagement in daily life and social interactions.
5. **Coping Strategies:** An individual's psychological state influences how they cope with tinnitus. People with healthy coping strategies and a positive outlook may be better equipped to manage the emotional impact of tinnitus.
6. **Cyclical Relationship:** Tinnitus and psychological distress can create a cyclical relationship. The distress caused by tinnitus can lead to increased focus on the sound, which in turn can worsen emotional distress. This cycle can be difficult to break without appropriate coping strategies.
7. **Quality of Life:** The emotional and psychological impact of tinnitus can significantly affect an individual's overall quality of life. Relationships, work, social activities, and daily functioning can all be compromised.
8. **Psychological Interventions:** Psychological interventions, such as cognitive behavioral therapy (CBT) and counseling, are often used to help individuals manage the emotional and psychological impact of tinnitus. These approaches can help individuals reframe their thoughts and develop effective coping strategies.
9. **Mindfulness and Relaxation Techniques:** Practices like mindfulness meditation and relaxation techniques can help reduce stress and improve emotional well-being, which can positively influence how a person perceives and copes with tinnitus.
10. **Support Networks:** Connecting with others who also experience tinnitus through support groups or online communities can provide emotional support and a sense of understanding, helping to reduce feelings of isolation.

Overall, the relationship between tinnitus and psychological state is complex and multidirectional. Addressing the emotional impact of tinnitus through psychological interventions, support networks, and lifestyle adjustments can be an important aspect of comprehensive tinnitus management. If you're experiencing emotional distress due to tinnitus, seeking help from mental health professionals and audiologists who specialize in tinnitus management can be beneficial.

**9.**

What are the best questionnaires to evaluate tinnitus?

ChatGPT

There are several questionnaires commonly used to evaluate the impact of tinnitus on individuals' lives. The choice of questionnaire depends on the specific goals of assessment, such as measuring the severity of tinnitus, its impact on daily functioning, emotional distress, and quality of life. Here are some of the most widely recognized questionnaires for evaluating tinnitus:

1. **Tinnitus Handicap Inventory (THI):** The THI is one of the most commonly used questionnaires to assess the impact of tinnitus on an individual's daily life. It consists of 25 items that cover emotional, functional, and catastrophic effects of tinnitus. The responses are scored to determine the degree of tinnitus-related handicap.
2. **Tinnitus Functional Index (TFI):** The TFI is a comprehensive questionnaire that assesses the functional and emotional impact of tinnitus. It includes 25 items that cover multiple domains, including intrusiveness, sense of control, cognitive interference, sleep disturbance, auditory perception, and quality of life.
3. **Tinnitus Questionnaire (TQ):** The TQ evaluates the psychological and emotional distress caused by tinnitus. It consists of 52 items that cover areas such as emotional and cognitive distress, sleep disturbance, auditory perception, and somatic complaints.
4. **Tinnitus Severity Index (TSI):** The TSI focuses specifically on the perceived severity of tinnitus. It involves rating the loudness and annoyance of tinnitus, as well as its effect on daily activities and sleep.
5. **Tinnitus Handicap Questionnaire (THQ):** The THQ assesses tinnitus-related handicap across different domains and can be used to track changes over time.
6. **Mini-Tinnitus Questionnaire (Mini-TQ):** The Mini-TQ is a shorter version of the TQ, focusing on the emotional distress and cognitive aspects of tinnitus.
7. **Goebel and Hiller Questionnaire:** This questionnaire measures tinnitus-related distress and can be particularly useful for assessing the psychological impact of tinnitus.
8. **Tinnitus Catastrophizing Scale (TCS):** The TCS assesses the degree to which individuals catastrophize about their tinnitus. It focuses on negative thinking patterns and emotional responses to tinnitus.
9. **Tinnitus Handicap Inventory-Screening Version (THI-S):** This shortened version of the THI is used as a screening tool to quickly assess the impact of tinnitus on daily life.

When selecting a questionnaire, consider the specific objectives of your assessment and the available resources. Some questionnaires are more comprehensive and may be better suited for detailed evaluations, while others are shorter and can serve as quick screening tools. It's important to note that while these questionnaires can provide valuable insights into the impact of tinnitus, they should be used in conjunction with clinical assessments and other diagnostic tools to form a comprehensive understanding of the condition.

**10.**

What is the expected result of auditory brainstem response in case of acoustic neuroma?

ChatGPT

Acoustic neuroma, also known as vestibular schwannoma, is a benign tumor that arises from the Schwann cells of the vestibulocochlear nerve (cranial nerve VIII). This nerve is responsible for transmitting auditory and balance information from the inner ear to the brainstem. The presence of an acoustic neuroma can potentially impact the auditory pathway, including the brainstem, and may lead to changes in the Auditory Brainstem Responses (ABRs).

In the context of an acoustic neuroma, the expected results of auditory brainstem responses can vary depending on factors such as the size of the tumor, its location, and the extent of its impact on the auditory pathway. Here are a few scenarios:

1. **Unilateral Hearing Loss:** Acoustic neuromas often cause unilateral (one-sided) hearing loss. In this case, ABRs recorded from the ear on the side of the tumor may show delayed or absent responses, particularly for higher-intensity or higher-frequency stimuli. This is due to the tumor's interference with the transmission of auditory signals from the cochlea to the brainstem.
2. **Waveform Abnormalities:** ABR waveforms are composed of several identifiable components (waves I, II, III, IV, and V), each representing neural activity at different levels of the auditory pathway. In the presence of an acoustic neuroma, the ABR waveform from the affected ear might show abnormalities, such as prolonged latencies or reduced amplitudes of specific waves. These abnormalities can provide insights into the tumor's impact on the auditory nerve and brainstem.
3. **Bilateral ABR Changes:** In some cases, an acoustic neuroma might affect both ears due to its location or growth pattern. Bilateral changes in ABR responses might indicate involvement of the auditory nerve on both sides.
4. **Asymmetry:** Comparing ABR responses between the ears can reveal asymmetry that could suggest the presence of a tumor. Significant asymmetry in ABR parameters may prompt further diagnostic investigations.

It's important to note that while ABRs can provide valuable information, they are just one part of a comprehensive diagnostic evaluation for acoustic neuroma. Imaging techniques such as MRI (magnetic resonance imaging) are essential for confirming the presence and location of the tumor.

If there is a suspicion of an acoustic neuroma or any other auditory-related condition, a thorough evaluation by an otolaryngologist (ear, nose, and throat specialist) and an audiologist is crucial. They can determine the appropriate diagnostic tests and treatment options based on the individual's specific situation.
