## Supplementary File 2 for "ChatGPT for tinnitus information and support: response accuracy and retest after three months"

Responses by ChatGPT – 26.11.2023

**1.**

I started to experience some strange sounds in the ear? What is it and what should I do?

ChatGPT

Experiencing strange sounds in the ear, such as ringing, buzzing, hissing, or other noises, is a common symptom known as tinnitus. Tinnitus is not a condition itself but rather a symptom of an underlying issue. While it can be bothersome, it is often not a sign of a serious medical problem. However, if you are experiencing persistent or bothersome tinnitus, it's essential to consult with a healthcare professional. Here are some steps you can take:

1. **Consult with an Audiologist or ENT Specialist:**
   - Schedule an appointment with an audiologist or an ear, nose, and throat (ENT) specialist to have your hearing and ear health assessed.
   - A comprehensive evaluation will help determine the cause and severity of your tinnitus.
2. **Identify Underlying Causes:**
   - Tinnitus can be caused by various factors, including exposure to loud noise, age-related hearing loss, earwax blockage, certain medications, or underlying health conditions.
   - Discuss your medical history and any recent changes or exposures with the healthcare professional.
3. **Hearing Evaluation:**
   - A hearing test (audiogram) may be conducted to assess your hearing abilities and identify any hearing loss that may be associated with tinnitus.
4. **Imaging Tests:**
   - In some cases, imaging tests like an MRI or CT scan may be recommended to rule out structural issues in the ear or head.
5. **Management and Treatment Options:**
   - Once the cause of tinnitus is identified, the healthcare professional will discuss appropriate management or treatment options.
   - Treatment may include addressing underlying medical conditions, using hearing aids for associated hearing loss, or employing strategies to manage and cope with tinnitus.
6. **Lifestyle Changes:**
   - Certain lifestyle changes may help alleviate tinnitus symptoms. These include reducing exposure to loud noises, managing stress, getting adequate sleep, and avoiding excessive use of headphones or earbuds.
7. **Sound Therapy:**
   - Sound therapy involves using external sounds, such as white noise or nature sounds, to help mask or distract from the perception of tinnitus. This can be achieved through wearable devices or smartphone apps.
8. **Counseling and Support:**
   - Counseling and support from healthcare professionals or support groups can be beneficial, especially if tinnitus is causing distress or impacting your quality of life.

Remember, it's crucial to consult with a healthcare professional for a proper diagnosis and guidance tailored to your specific situation. While tinnitus may not always have a cure, many people find relief through various management strategies. If you notice a sudden onset of tinnitus, especially if it's accompanied by hearing loss or dizziness, seek medical attention promptly.

**2.**

How can I help myself when I suffer from tinnitus?

ChatGPT

While there may not be a cure for tinnitus, there are several strategies and lifestyle changes that individuals with tinnitus can adopt to manage and alleviate the symptoms. Here are some self-help tips for coping with tinnitus:

1. **Consult with Healthcare Professionals:**
   - Seek guidance from an audiologist or an ear, nose, and throat (ENT) specialist to identify the underlying cause of your tinnitus and receive appropriate treatment.
2. **Manage Stress:**
   - Stress and anxiety can exacerbate tinnitus symptoms. Practice stress-reducing techniques such as deep breathing, meditation, yoga, or progressive muscle relaxation.
3. **Sound Therapy:**
   - Use external sounds to mask or distract from the tinnitus. White noise machines, fans, or soothing music can be helpful, especially in quiet environments.
4. **Hearing Aids:**
   - If you have hearing loss along with tinnitus, using hearing aids may help by amplifying external sounds and providing relief.
5. **Avoid Loud Noises:**
   - Protect your ears from loud noises, as exposure to loud sounds can worsen tinnitus. Use earplugs or noise-canceling headphones in noisy environments.
6. **Limit Caffeine and Nicotine:**
   - Caffeine and nicotine can contribute to tinnitus in some individuals. Consider reducing or eliminating your intake to see if it makes a difference.
7. **Regular Exercise:**
   - Engage in regular physical activity, as exercise can improve blood flow and reduce stress. Choose activities you enjoy, whether it's walking, swimming, or cycling.
8. **Healthy Sleep Habits:**
   - Establish a consistent sleep routine and create a comfortable sleep environment. Consider using white noise or other calming sounds to help you relax.
9. **Mindfulness and Cognitive Behavioral Therapy (CBT):**
   - Mindfulness techniques and CBT can be effective in managing the emotional aspects of tinnitus. These approaches help change negative thought patterns and reactions to tinnitus.
10. **Limit Exposure to Loud Music:**
    - When using headphones or earbuds, keep the volume at a moderate level to avoid further damage to your hearing.
11. **Support Groups:**
    - Joining a support group or connecting with others who experience tinnitus can provide valuable insights, coping strategies, and emotional support.
12. **Consult Alternative Therapies:**
    - Some people find relief through acupuncture, hypnosis, or other alternative therapies. While the effectiveness varies, these approaches may be worth exploring.
13. **Stay Informed:**
    - Learn more about tinnitus and its management. Understanding the condition can help you feel more in control and reduce anxiety.

Remember that what works for one person may not work for another, so it may take some time to find the strategies that work best for you. It's important to consult with healthcare professionals for personalized advice based on your specific situation. If your tinnitus is causing significant distress or if you experience sudden or severe symptoms, seek prompt medical attention.

**3.**

Should I believe in the advertisement of a medicine that treats tinnitus in one week?

ChatGPT

While advertisements for medications claiming to treat tinnitus quickly may be tempting, it's essential to approach such claims with caution and skepticism. Tinnitus is a complex symptom with various potential underlying causes, and there is currently no universally accepted, quick-fix cure for all cases. Here are some considerations to keep in mind:

1. **No Universal Cure:** Tinnitus is not a specific disease but a symptom of an underlying condition. The causes of tinnitus can vary, including age-related hearing loss, exposure to loud noise, earwax blockage, or underlying medical issues. A one-size-fits-all cure is unlikely.
2. **Individual Variability:** Tinnitus affects individuals differently. What works for one person may not work for another due to the diverse causes and contributing factors.
3. **Limited Scientific Evidence:** Be cautious if the advertisement lacks credible scientific evidence, such as clinical trials and peer-reviewed studies, supporting the effectiveness of the medication. Scientifically rigorous research is crucial in validating the claims of any medical treatment.
4. **Consult Healthcare Professionals:** Before trying any medication or treatment for tinnitus, it is advisable to consult with a healthcare professional, preferably an audiologist or an ear, nose, and throat (ENT) specialist. They can assess your specific case, identify the potential cause of tinnitus, and recommend appropriate treatments or management strategies.
5. **Beware of Misleading Claims:** Be wary of advertisements that make unrealistic promises or use sensational language. Claims of a "quick cure" or "guaranteed results" without proper scientific support are often red flags.
6. **Consider Lifestyle Factors:** Tinnitus management often involves addressing lifestyle factors, such as reducing exposure to loud noise, managing stress, and adopting healthy habits. Quick fixes may not address these underlying contributors.
7. **Holistic Approach:** A holistic approach to managing tinnitus may involve a combination of strategies, including sound therapy, counseling, and lifestyle modifications. Medications, if recommended, should be part of a comprehensive treatment plan.
8. **Patient Testimonials:** While patient testimonials may provide insights, they should not be the sole basis for making decisions. Individual responses to treatments can vary, and anecdotal evidence may not represent the broader population.

It's crucial to be discerning and prioritize evidence-based treatments when considering options for tinnitus management. If you encounter an advertisement making bold claims about curing tinnitus quickly, it's advisable to seek guidance from a qualified healthcare professional who can provide personalized advice based on a thorough evaluation of your condition.

**4.**

How to diagnose tinnitus?

ChatGPT

Diagnosing tinnitus involves a comprehensive evaluation by healthcare professionals to determine the underlying cause and assess the impact of the condition on an individual's hearing and quality of life. Here are the key steps in the diagnostic process for tinnitus:

1. **Medical History:**
   - The healthcare professional will begin by taking a detailed medical history, asking questions about the nature of the tinnitus, its onset, duration, and any associated symptoms. Information about past medical conditions, medications, and exposure to loud noise or ototoxic substances is also important.
2. **Physical Examination:**
   - A physical examination, including an examination of the head, neck, and ears, will be conducted to identify any visible abnormalities or signs of an underlying medical condition.
3. **Hearing Evaluation (Audiological Assessment):**
   - A hearing test, also known as an audiogram, is typically conducted to assess hearing abilities. This test helps identify any hearing loss and provides information about the type and degree of hearing impairment.
4. **Pitch and Loudness Matching:**
   - During an audiometric evaluation, individuals with tinnitus may undergo pitch and loudness matching tests. These tests help determine the frequency (pitch) and intensity (loudness) of the perceived tinnitus sound.
5. **Tinnitus Questionnaires:**
   - Healthcare professionals may use validated tinnitus questionnaires to gather information about the impact of tinnitus on an individual's daily life, emotional well-being, and overall quality of life.
6. **Imaging Studies:**
   - In some cases, imaging studies such as magnetic resonance imaging (MRI) or computed tomography (CT) scans may be recommended to rule out structural issues or abnormalities in the head and neck that could be contributing to tinnitus.
7. **Specialized Tests:**
   - Additional specialized tests may be conducted based on the suspected cause of tinnitus. These tests can include otoacoustic emissions (OAEs), auditory brainstem response (ABR) testing, and other electrophysiological tests.
8. **Consultation with Specialists:**
   - Depending on the findings, individuals with tinnitus may be referred to specialists, such as an audiologist, otolaryngologist (ENT), neurologist, or other healthcare professionals, for further evaluation and management.

It's important to note that tinnitus itself is a symptom and not a disease. The diagnostic process aims to identify any underlying conditions or contributing factors that may be causing or exacerbating the tinnitus. Once a thorough evaluation is completed, healthcare professionals can develop an appropriate treatment plan or recommend management strategies tailored to the individual's specific case. If you are experiencing tinnitus, it is advisable to consult with a healthcare professional for a proper diagnosis and guidance.

**5.**

Is there a connection between hearing loss and tinnitus?

ChatGPT

Yes, there is a strong connection between hearing loss and tinnitus. In fact, tinnitus is often associated with hearing loss, and the two conditions frequently coexist. Here's how they are connected:

1. **Common Underlying Causes:**
   - Hearing loss and tinnitus can share common underlying causes. One of the most common causes is damage to the hair cells in the inner ear, which can occur due to aging, exposure to loud noise, ototoxic medications, or other factors.
2. **Cochlear Damage:**
   - In many cases, both hearing loss and tinnitus result from damage to the cochlea, the sensory organ of hearing in the inner ear. This damage can lead to changes in the way the auditory system processes sound, contributing to the perception of both reduced hearing and the sensation of ringing or buzzing (tinnitus).
3. **Neural Compensation and Hyperactivity:**
   - When the auditory system experiences hearing loss, the brain may undergo changes in an attempt to compensate for the reduced input of auditory signals. This neural compensation can lead to increased neural activity, sometimes interpreted as the perception of sound, resulting in tinnitus.
4. **Frequency Matching:**
   - Tinnitus often occurs in conjunction with specific patterns of hearing loss. The pitch (frequency) of the perceived tinnitus sound may correspond to the frequencies affected by hearing loss. This relationship is identified through pitch matching during audiometric evaluations.
5. **Prevalence in Hearing Loss Population:**
   - Studies have shown that a significant percentage of individuals with hearing loss also experience tinnitus. The relationship is particularly pronounced in cases of sensorineural hearing loss, where the damage occurs in the inner ear or the auditory nerve.
6. **Treatment Considerations:**
   - Hearing aids are a common and effective treatment option for individuals with both hearing loss and tinnitus. By amplifying external sounds and improving overall auditory input, hearing aids can often help reduce the perception of tinnitus.
7. **Auditory Pathway Interactions:**
   - The interconnected nature of the auditory pathways means that changes in one part of the system can affect other components. Hearing loss can disrupt the normal functioning of these pathways, leading to alterations in neural activity that contribute to the development and persistence of tinnitus.

It's important to note that while hearing loss and tinnitus often coexist, not everyone with hearing loss experiences tinnitus, and vice versa. Additionally, the severity and impact of tinnitus can vary widely among individuals. If you are experiencing hearing loss or tinnitus, seeking the guidance of an audiologist or an ear, nose, and throat (ENT) specialist is recommended for a thorough evaluation and appropriate management strategies.

**6.**

What is the difference between objective and subjective tinnitus?

ChatGPT

The terms "objective tinnitus" and "subjective tinnitus" refer to different types of tinnitus based on the perception of the sound and the ability to measure it. Here's the difference between the two:

1. **Objective Tinnitus:**
   - **Definition:** Objective tinnitus is characterized by sounds that can be heard not only by the person experiencing tinnitus but also by others, including healthcare professionals during an examination.
   - **Causes:** Objective tinnitus is often associated with physical sounds generated by the body, such as blood flow, muscle spasms, or abnormalities in the middle ear. These sounds can be audible using specialized instruments during a clinical examination.
   - **Examples:** Pulsatile tinnitus is a common form of objective tinnitus where the person perceives a rhythmic sound that coincides with the heartbeat. This can be due to blood flow turbulence.
2. **Subjective Tinnitus:**
   - **Definition:** Subjective tinnitus refers to sounds that are perceived only by the individual experiencing tinnitus. These sounds are not audible to others and cannot be measured externally.
   - **Causes:** Subjective tinnitus is often associated with damage to the auditory system, particularly the inner ear (cochlea) or the auditory nerve. It can also result from conditions such as age-related hearing loss, exposure to loud noise, or certain medical conditions.
   - **Examples:** Ringing, buzzing, hissing, or other sounds that the individual hears without an external source are common examples of subjective tinnitus.

In summary, the key difference lies in whether the tinnitus sounds can be objectively measured or heard by others. Objective tinnitus involves sounds that can be detected externally, often due to physical processes in the body, while subjective tinnitus involves sounds that are perceived only by the individual and cannot be measured externally. Subjective tinnitus is far more common and represents the majority of tinnitus cases. If you are experiencing tinnitus, it's important to consult with a healthcare professional, such as an audiologist or an ear, nose, and throat (ENT) specialist, for a comprehensive evaluation and appropriate management.

**7.**

How the tinnitus is connected to otoacoustic emissions?

ChatGPT

The connection between tinnitus and otoacoustic emissions (OAEs) lies in the underlying mechanisms of the auditory system and how disturbances in these mechanisms can contribute to the perception of tinnitus. OAEs are sounds generated by the inner ear in response to external stimuli, and they play a role in our understanding of both normal auditory function and certain hearing disorders, including tinnitus. Here's how they are connected:

1. **Cochlear Function:**
   - OAEs are sounds produced by the cochlea, the spiral-shaped organ in the inner ear responsible for converting sound vibrations into electrical signals that can be interpreted by the brain. Disturbances in cochlear function can lead to changes in OAE responses.
2. **Role of OAEs in Tinnitus Research:**
   - Researchers use OAEs as a tool to study the health and function of the cochlea. Changes in OAE responses may indicate abnormalities or damage to the cochlea, providing insights into the mechanisms underlying tinnitus.
3. **Associations with Hearing Loss:**
   - OAEs are often used to assess hearing function, and disruptions in OAEs are associated with certain types of hearing loss. Since tinnitus is commonly linked to hearing loss, particularly sensorineural hearing loss, abnormalities in OAEs can be observed in individuals with tinnitus.
4. **Hidden Hearing Loss:**
   - Hidden hearing loss refers to difficulties in hearing that are not always detected by standard audiometric tests. OAEs are sensitive to changes in cochlear function, making them useful in identifying hidden hearing loss, which may be associated with tinnitus.
5. **Auditory Neuropathy Spectrum Disorder (ANSD):**
   - OAEs are also valuable in the diagnosis of auditory neuropathy spectrum disorder (ANSD), a condition where the transmission of signals from the inner ear to the auditory nerve is impaired. ANSD can be associated with tinnitus.
6. **Correlation with Tinnitus Severity:**
   - Some studies have explored the correlation between OAE responses and the severity of tinnitus. While not everyone with tinnitus will show abnormalities in OAEs, changes in OAEs may be more common in individuals with more severe or persistent tinnitus.
7. **Clinical Applications:**
   - OAE measurements are used clinically to assess cochlear function, especially in cases where hearing loss is suspected. This information can be relevant to understanding the connection between cochlear health, hearing loss, and tinnitus.

It's important to note that while OAEs can provide valuable information about the health of the auditory system and its role in hearing loss and tinnitus, they are not a direct measure of tinnitus itself. Tinnitus is a complex symptom influenced by various factors, including neural processing, psychological factors, and individual differences. If you are experiencing tinnitus, a comprehensive evaluation by a healthcare professional, including assessments like OAEs and audiometry, can help identify potential contributing factors and guide appropriate management strategies.

**8.**

Is there link between tinnitus and psychological state of the patient?

ChatGPT

Yes, there is a well-established link between tinnitus and the psychological state of the patient. Tinnitus is not solely an auditory phenomenon but also involves complex interactions between the auditory system, the brain, and psychological factors. Here are key aspects of the connection between tinnitus and psychological well-being:

1. **Impact on Emotional Health:**
   - Tinnitus can have a significant impact on emotional well-being. The persistent perception of sound, especially when it is bothersome or disruptive, can lead to emotional distress, anxiety, and depression.
2. **Stress and Anxiety:**
   - The presence of tinnitus can contribute to increased stress levels and anxiety. Individuals with tinnitus may experience heightened awareness of the sound, leading to a cycle of stress and exacerbation of tinnitus symptoms.
3. **Quality of Life:**
   - Tinnitus can affect an individual's overall quality of life. The perceived intrusiveness of the sound, along with its potential impact on concentration, sleep, and daily activities, can contribute to a diminished quality of life.
4. **Sleep Disturbances:**
   - Many individuals with tinnitus report difficulties with sleep. The constant background noise of tinnitus may interfere with falling asleep or staying asleep, leading to fatigue and further contributing to emotional distress.
5. **Negative Thought Patterns:**
   - Tinnitus can give rise to negative thought patterns, including feelings of frustration, helplessness, or a sense of loss. These thoughts can exacerbate emotional distress and create a negative feedback loop.
6. **Cognitive Factors:**
   - The cognitive aspects of tinnitus, such as the perception of its loudness or the degree of distress it causes, are influenced by psychological factors. Individual differences in coping mechanisms and resilience play a role in how tinnitus is experienced.
7. **Psychosomatic Interactions:**
   - Tinnitus can interact with psychosomatic processes, where psychological factors may influence the perception and severity of tinnitus symptoms. Conversely, the distress caused by tinnitus can manifest in physical symptoms.
8. **Counseling and Therapy:**
   - Psychological interventions, such as counseling and cognitive-behavioral therapy (CBT), are commonly used in the management of tinnitus. These approaches aim to address the emotional and cognitive aspects of tinnitus and provide coping strategies.
9. **Support Groups:**
   - Support groups and peer interactions can be beneficial for individuals with tinnitus. Sharing experiences and coping strategies with others who have tinnitus can help reduce feelings of isolation and provide emotional support.

Understanding the psychological aspects of tinnitus is crucial for developing comprehensive management strategies. A multidisciplinary approach that includes both audiological and psychological components is often effective in addressing the diverse aspects of tinnitus and improving the overall well-being of individuals with tinnitus. If you or someone you know is experiencing distress due to tinnitus, seeking the guidance of healthcare professionals, including audiologists, psychologists, or counselors, is recommended.

**9.**

What are the best questionnaires to evaluate tinnitus?

ChatGPT

Several validated questionnaires are commonly used to evaluate the impact of tinnitus on individuals. These questionnaires help assess the severity of tinnitus symptoms, the degree of distress it causes, and its impact on various aspects of daily life. Here are some widely used tinnitus questionnaires:

1. **Tinnitus Handicap Inventory (THI):**
   - **Purpose:** The THI assesses the impact of tinnitus on daily life and categorizes it into functional, emotional, and catastrophic aspects.
   - **Content:** The questionnaire consists of 25 items, with respondents indicating the degree of impact on each aspect.
   - **Scoring:** Scores range from 0 to 100, with higher scores indicating greater perceived handicap.
2. **Tinnitus Functional Index (TFI):**
   - **Purpose:** The TFI is a comprehensive questionnaire that assesses the functional, emotional, and catastrophic impact of tinnitus.
   - **Content:** It includes 25 items covering various aspects, including intrusiveness, sense of control, sleep, and cognitive interference.
   - **Scoring:** Scores range from 0 to 100, with higher scores indicating greater functional impairment.
3. **Tinnitus Reaction Questionnaire (TRQ):**
   - **Purpose:** The TRQ assesses the emotional and cognitive reactions to tinnitus.
   - **Content:** It includes 27 items that measure the degree of emotional distress and cognitive difficulties related to tinnitus.
   - **Scoring:** Scores range from 0 to 27, with higher scores indicating greater emotional and cognitive reactions.
4. **Tinnitus Severity Index (TSI):**
   - **Purpose:** The TSI assesses the severity of tinnitus symptoms across different domains.
   - **Content:** It includes 12 items covering loudness, awareness, annoyance, and emotional reactions to tinnitus.
   - **Scoring:** Scores range from 0 to 100, with higher scores indicating greater severity.
5. **Tinnitus Handicap Questionnaire (THQ):**
   - **Purpose:** The THQ evaluates the impact of tinnitus on daily life and emotional well-being.
   - **Content:** It includes 13 items that assess the emotional, social, and occupational aspects of tinnitus impact.
   - **Scoring:** Scores range from 0 to 100, with higher scores indicating greater handicap.
6. **Tinnitus Primary Function Questionnaire (TPFQ):**
   - **Purpose:** The TPFQ assesses the primary functions affected by tinnitus, including hearing, concentration, and sleep.
   - **Content:** It includes 18 items, and respondents rate the impact of tinnitus on various functions.
   - **Scoring:** Scores range from 0 to 54, with higher scores indicating greater impact on primary functions.

When using these questionnaires, it's important to consider the specific goals of the assessment and choose the one that aligns with the aspects of tinnitus impact you are interested in measuring. Additionally, a healthcare professional, such as an audiologist or psychologist, can help interpret the questionnaire results and guide appropriate management strategies based on the individual's tinnitus experience.

**10.**

What is the expected result of auditory brainstem response in case of acoustic neuroma?

ChatGPT

Auditory Brainstem Response (ABR) testing is a diagnostic procedure used to assess the integrity of the auditory nerve and the auditory pathways in the brainstem. In the case of an acoustic neuroma, which is a benign tumor that affects the vestibulocochlear nerve (the eighth cranial nerve responsible for hearing and balance), ABR results can show specific patterns indicative of the tumor's presence. However, it's important to note that the ABR alone cannot definitively diagnose an acoustic neuroma, and additional imaging studies such as magnetic resonance imaging (MRI) are typically required for confirmation.

Here are the expected ABR results in the context of an acoustic neuroma:

1. **Delayed Wave I Latency:**
   - In the presence of an acoustic neuroma, ABR results may show a delayed Wave I latency. Wave I represents the electrical activity generated by the auditory nerve in response to a sound stimulus. The delay is attributed to the tumor's impact on the normal transmission of signals along the auditory nerve.
2. **Increased Interpeak Latencies:**
   - Interpeak latencies represent the time intervals between different waves in the ABR waveform. The presence of an acoustic neuroma can lead to increased interpeak latencies, indicating disruptions in the normal transmission of auditory signals along the auditory pathway.
3. **Wave V Absence or Delay:**
   - Wave V is a prominent component of the ABR waveform, representing activity in the brainstem. In cases of acoustic neuroma, Wave V may be absent or delayed due to the tumor's interference with the auditory pathways.

It's important to emphasize that while ABR findings can raise suspicion of an acoustic neuroma, the definitive diagnosis is typically confirmed through imaging studies, especially high-resolution MRI. MRI can provide detailed images of the internal auditory canal and identify the presence, size, and location of the tumor.

If an individual is suspected of having an acoustic neuroma due to abnormal ABR findings, further evaluation and management by an otolaryngologist (ENT specialist) and a neurosurgeon are essential. Early detection and intervention are crucial for managing acoustic neuromas and preserving hearing function.

It's also worth noting that ABR is a valuable tool in various clinical contexts beyond acoustic neuroma diagnosis, such as assessing hearing thresholds, identifying auditory nerve abnormalities, and monitoring auditory function during surgeries.
